# A new method for noninvasive determination of effective pulmonary blood flow (EPBF) and cardiac output (CO) in spontaneously breathing subjects

**DOI:** 10.1101/2025.06.26.25330339

**Authors:** Andras Gedeon, Jakob Jansson, David Patriksson, Mats Wallin

## Abstract

The differential Fick method is well established for measuring EPBF and CO but until now it has only been used for patients on mechanical ventilation. Here we present and evaluate a new approach adapted to spontaneous breathing situations.

Ten healthy subjects with diverse anthropometric and respiratory parameters were studied in the sitting position. Rebreathing through dead space of precisely known volume and recording the resulting rise in the end-tidal CO_2_ value allows the determination of EPBF. The shunted blood flow fraction is estimated from the arterial oxygen saturation to obtain cardiac output (FickCO). Two measurements were made on each subject 15 minutes apart. Reference values for cardiac output (RefCO), were calculated as the product of stroke volume and heart rate where the stroke volume was measured with established echocardiography techniques. Heart rate and arterial oxygen saturation were measured with an ordinary pulse oximeter

Comparing FickCO to Ref CO using the Bland-Altman analysis we obtain bias = 0.03 l/min and SD = 0.71 l/min resulting in a percentage error PE = 0.25

The differential Fick method can be adapted to spontaneously breathing situations with good absolute accuracy using simple equipment. Short data collection times make it possible to use the mean of repeated observations and thereby get adequate precision. The new method could therefore be of value both in the pre-operative and the post-operative setting.

## Introduction

The differential Fick method, also called indirect Fick, is well established for the measurement of non-shunted pulmonary blood flow (EPBF). [1–4]. The method utilizes the fact that due to the large body stores of carbon dioxide a suddenly induced change in the CO_2_ elimination (VCO_2_) from the lungs will immediately change the arterial blood content while the mixed venous blood content will remain unchanged for a certain period of time. During this time interval the change in VCO_2_ will be proportional to the change in the arterial CO_2_ partial pressure which in turn can be approximated with the change in the end tidal CO_2_ (Pet). The procedure is non-invasive, and the constant of proportionality determines EPBF.

There are several ways to change VCO_2,_ and it is most conveniently done when the patient is on a mechanical ventilator. Therefore, till now, the method has been used only for ventilated patients in the ICU or OR. Initial work [1] as well as the most recent developments [5–7] have used manipulations of the breathing pattern of the patient. However, since the first introduction of the CO_2_ rebreathing approach [8] this way to change VCO_2_ has been well studied [2,4,9,10] and also the method chosen for a commercial implementation [4].

In the present work we adopt the rebreathing method and extend its use to measure EPBF also in spontaneously breathing subjects and we evaluate how well the system performs relative to cardiac output measurements made by standard echocardiography.

## Methods

Ten subjects, 4 women and 6 men, with no known cardio-pulmonary disease, were recruited and studied in the sitting position. They were between 39-70 (mean 55) years old, with weights between 62-99 (mean 80) kg, tidal volume (TV) was between 0.58-1.70 (mean 1.1) l and respiratory rate (RR) between 4.6-16.8 (mean 11) breath/minute.

The equipment used for this study is shown in Fig 1. It is made up of a nose clip, a dead space volume (DV), and a sampling IR CO2 analyzer, ET600 Side stream CO2CGM Module (Ronseda Electronics Co. Ltd China) and a laptop computer for data collection. To assess pulmonary function, TV and functional residual capacity (FRC) were measured with Cosmed Quark CPET (COSMED SRl Italy).

**Fig 1.**
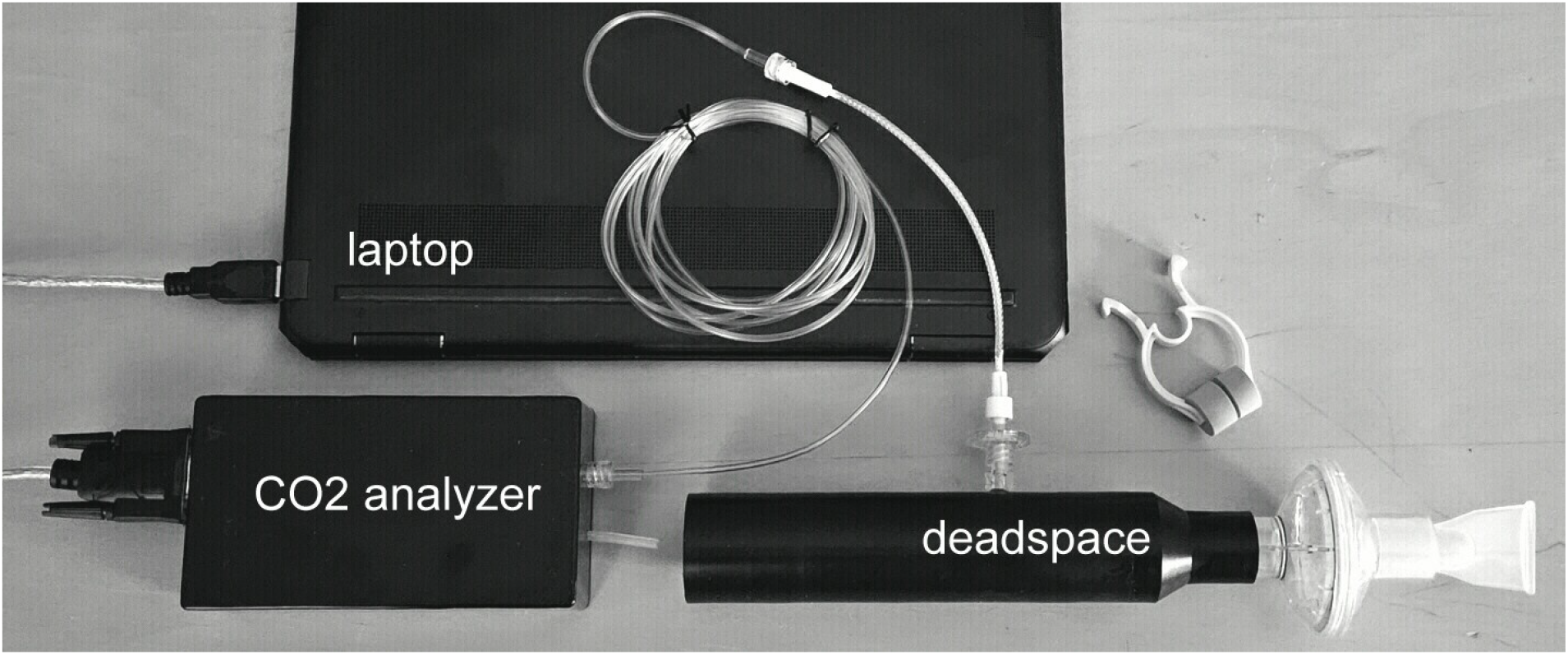

The dead space volume is formed by connecting together a mouthpiece, a respiratory filter (ZF-007 Shandong Zhenfu Medical Device Co., Ltd China) and a cylindrical pipe. The combined volume is determined with an accuracy of better than 3 ml. The sampling port of the analyzer is connected to the pipe through a nafion tube and a sampling filter. The sampling point is ~10 mm inside the lumen of the pipe. Three different volumes were used: DV=0.168 l, DV=0.238 l and DV=0.306 l. They were chosen so that the ratio DV/TV was in the range ~ 0.2-0.3.

The subjects were instructed to sit still for about 5 minutes. During this time a pulse oximeter (Braun Pulse Oximeter 1) was applied to a finger and the heart rate (HR) and the oxygen saturation SaO_2_ were noted. We observed SaO_2_ in the range 93-99 (mean 96) % and HR in the range 60-86 (mean 73) beats/min. The subjects were then asked to apply the nose clip and to start breathing in the dead space, beginning with an expiration and to breathe in a normal manner till instructed to stop. Data collection took typically 30 seconds and always less than 50 seconds. After sitting still for about 15 minutes this procedure was repeated for a second set of measurements.

Reference cardiac output (RefCO) was measured in the sitting position with a GE Vivid E95 ultrasound unit using the VTI-method. Here the stroke volume is obtained from the approximately circular area of outflow from the left ventricle, times the outflow velocity of the blood. As a validation check, the stroke volume was also calculated from the 3D reconstruction of the left ventricle as the difference between the volumes pre and post systole. The two methods agreed within about 3% in all cases.

According to the differential Fick method [1] the effective pulmonary blood flow, EPBF can be obtained from the expression:

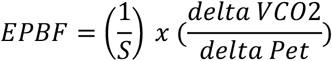

where S is the slope of the CO_2_ dissociation curve, delta VCO2 is the reduction in CO_2_ outflow from the lungs due to rebreathing and delta Pet is the resulting increase in the end-tidal Pet value.

In our approach [12], deltaVCO_2_ is obtained by first determining the volume of CO_2_ within the dead space at the end of expiration and then by multiplying this value with the mean respiratory rate, meanRR during rebreathing. The CO_2_ volume in the dead space is calculated as the product of the known dead space volume and a representative sample of the CO_2_ partial pressure within the dead space at the end of expiration.

With DV/TV < 0.3, the CO_2_ partial pressure will decrease slowly and linearly within the dead space away from the subject. A sampling point that divides the dead space into two equal parts will therefore provide a representative average value for the entire dead space, avPet. We thus get

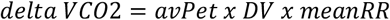

We use Capek’s formula for S [11]

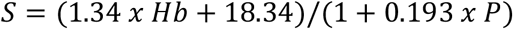

with Hb = 13.8 for women and Hb=15.0 for men. In accordance with previous analysis [1,9] for healthy subjects the slope can be evaluated at a CO_2_ partial pressure P = Pet + 10 mmHg where Pet is value measured at the start of rebreathing.

Shunted blood flow, passing where the ventilation/perfusion ratio in the lung is small or zero, can be estimated from SaO_2_ values based on the work of Nunn [13]. His results have been given a graphical presentation [14] and from this an analytical expression can be found for the shunt fraction (Sh) as a function of SaO_2_. For breathing room air, we get

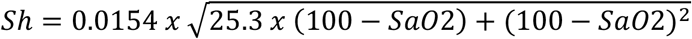

The cardiac output from our measurements, FickCO, to be compared to the reference, RefCO is then given by

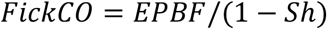

## Results

Fig 2 shows two cases of raising Pet due to rebreathing. On the left a nearly perfect data set and on the right a more typical recording. The left recording produces a well-defined delta Pet with the result FickCO = 6.1 l/min to be compared with RefCO = 5.9 l/min. The recording on the right opens for alternative analyses. Two different possible delta Pet values are shown. They correspond to FickCO = 5.2 l/min and FickCO = 5.7 l/min respectively that is an uncertainty of 0.5 l/min or about 10%. In this case Ref CO = 5.3 l/min indicating that delta2 Pet is the more correct interpretation.

**Fig 2.**
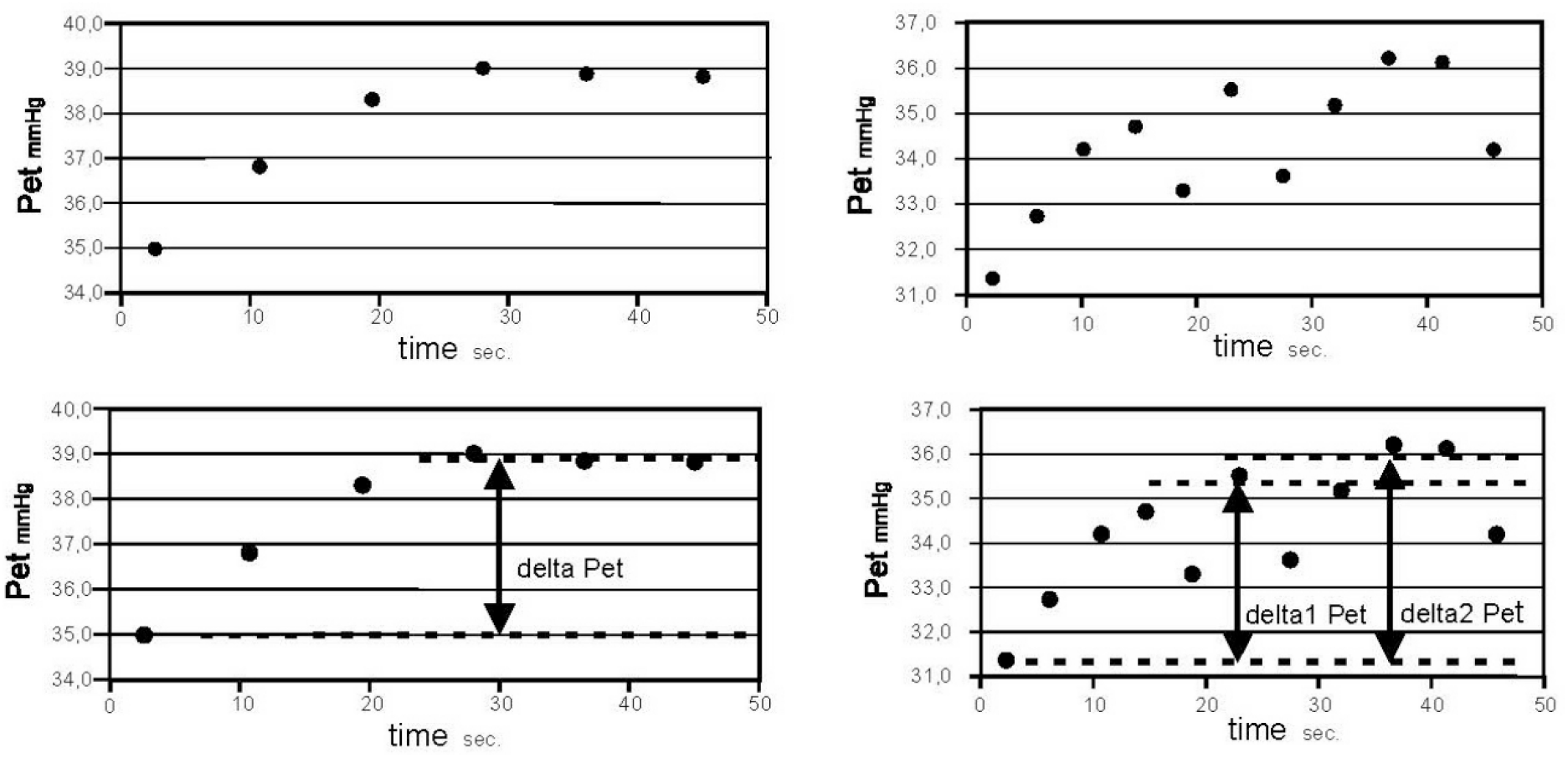

We were unable to reliably evaluate four registrations in the first set of measurements and three in the second due to irregular breathing and/or leakage around the mouthpiece. We thus obtained 13 data points for the comparison to RefCO. The result is shown in Fig 3 using the Bland-Altman presentation [15] for the RefCO vs FickCO data. We obtain a bias = 0.03 l/min with a SD = 0.71 l/min. and a percentage error PE= 0.25

**Fig 3.**
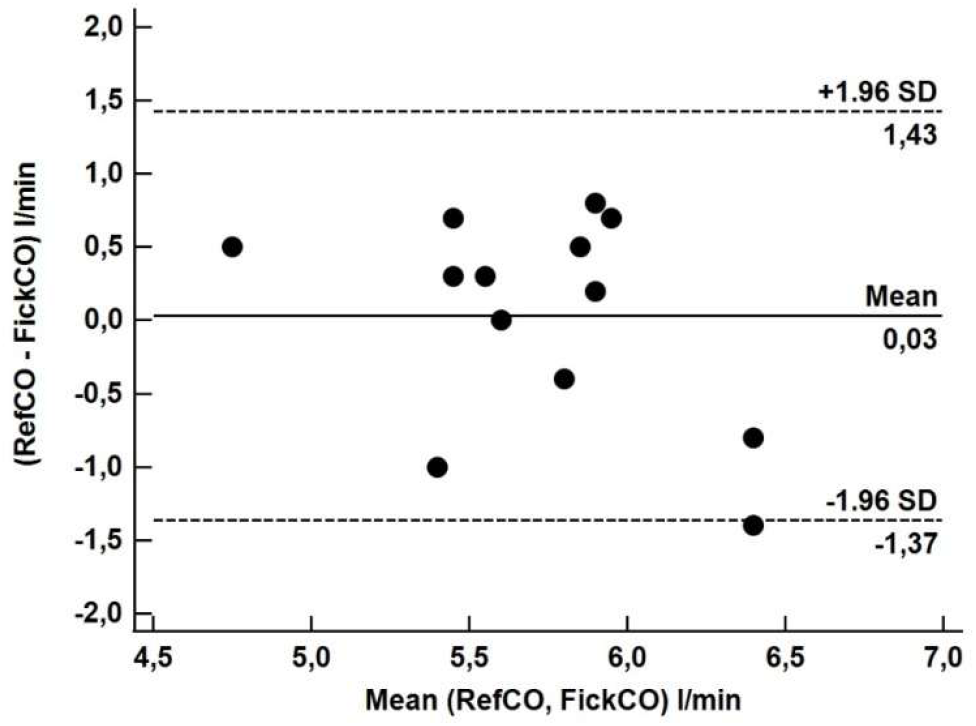
shows the Bland-Altman presentation for the RefCO vs FickCO data.

## Discussion

For spontaneous breathing situations, rebreathing is a natural choice for altering VCO_2_. It perturbs the alveolar ventilation without involving cooperation from the subject and allows for a very simple measuring system that is in addition highly fault tolerant [8,12]. The expression for EPBF is a ratio of two differences and since all data are collected with a single analyzer the result will be insensitive both to instrument off-set and to the gain setting.

Furthermore, by obtaining VCO_2_ using precise volume measurements and not as is usual from integrated flow and concentration vs time data, our approach offers both simplicity and better measuring accuracy. When comparing our method to the reference method, the exchangeability criteria of Chrichley [16] is fulfilled.

The dead space volume chosen should not be too small compared to the tidal volume because this would produce smaller increases in Pet and reduce useful information and not too large because this might lead to a non-linear concentration gradient within the dead space so that the Pet value might no longer be representative of the mean concentration in the entire volume. We have found that DV/TV = 0.2-0.3 is a suitable interval for measuring EPBF.

The slope of the CO_2_ dissociation curve (S) changes slowly and although it depends on parameters that are not exactly known, they can still be estimated well enough not to give rise to significant errors. As an example, the CO_2_ partial pressure where the slope of the CO_2_ dissociation curve (S) is to be determined depends on the end-tidal to arterial PCO_2_ difference. In our healthy subjects this is typically on average 4 mmHg [17]. A change of ~ 2 mmHg in this value would change FickCO about 5%. Also, if Hb in the expression for S was to change 10% this would also change FickCO about 5%. In both cases it would change the bias when compared to RefCO with about 0.25 l/min.

The shunt fraction calculation used to obtain FickCO from EPBF introduces both absolute error and variability. A typical error of +-2% in the value of SaO_2_ translates into an error in FickCO of about +-9% when SaO_2_= 98% while the same error in SaO_2_ translates into an error of only about +-5.5% when SaO_2_= 96%.

The most challenging aspect of the new method contributing to the variability in the data is irregular breathing. As shown in Fig 2 in a typical case, this can be about 10%. Finding ways to promote regular breathing is therefore of essence. In any case, precision can be improved by repeating the measurement and using average values instead of single observations. In the present study we have chosen to repeat the measurement once, after 15 minutes, to ascertain that the gas exchange changes introduced by the first measurement had no influence on the second.

However, there are good reasons to believe that with a typical rebreathing time of only about 30 seconds the measurement could have been repeated faster, possibly as often as every 5 minutes [11].

## Conclusions

The differential Fick method can be adapted to spontaneous breathing situations using simple equipment. The absolute accuracy of FickCO compares favorably with the clinically accepted reference. Short measuring times allow for using an average of repeated measurements assuring satisfactory precision. The method could have potentially considerable value both in the pre-operative and post-operative settings. Attention should be given to setting up test conditions and procedures that favor regular breathing.

## Data Availability

All data produced in the present work are contained in the manuscript.

## Acknowledgements

Dr R Winter for placing the resources of eHeart AB to our disposal. Clinical Trial Consultants AB (Uppsala Sweden) for recruiting the study subjects.

